# Women-related cardiovascular research funding under Canada’s Sex- and Gender-Based Analysis policy from 2000 to 2024: an interrupted time series analysis

**DOI:** 10.64898/2026.01.14.26344152

**Authors:** Nan Chen, Tamil Kendall, Wei Zhang, Lori A. Brotto

## Abstract

**Background:** Cardiovascular disease is the leading cause of death among women, yet women have historically been underrepresented in cardiovascular research. In Canada, sex- and gender-based analysis (SGBA) policies were introduced to address these gaps, including mandatory applicant-level requirements in 2010 and reviewer-level guidance in 2018. How these policies have influenced funding allocation for women-related cardiovascular research remains unclear.

**Objective:** To examine long-term trends in CIHR investment in women-related cardiovascular research and assess changes associated with SGBA policy milestones.

**Methods:** We conducted a longitudinal study using Canadian Institutes of Health Research (CIHR) funding data from fiscal year 2000-2001 to 2024-2025. Women-related cardiovascular research projects were identified through terminology searches in funded project titles, keywords, and abstracts. Annual fiscal-year proportions of cardiovascular research funding allocated to women-related research were analysed using segmented regression, with interruption points in 2010 and 2018.

**Results:** Among 17,168 cardiovascular research related grant annual records, 11.33% were classified as women-related projects. These projects accounted for 11.85% of total CIHR cardiovascular research funding over the 25-year period. Funding increased before 2010, declined between 2011 and 2017, and accelerated after 2018. Segmented regression showed a small immediate increase in 2010, followed by a negative post-2010 trend and a significant positive quadratic trend after 2018.

**Conclusions:** Application-level SGBA requirements had limited influence on investment patterns, whereas the 2018 reviewer-level guidance aligns with the subsequent acceleration in women-related cardiovascular research funding. Strengthening SGBA implementation, expanding targeted funding opportunities, and improving monitoring of sex-and-gender-disaggregated outputs may help address persistent gaps in women-related cardiovascular research.

## Background

Cardiovascular disease is the leading cause of death among women, accounting for approximately 35% of all female deaths worldwide in 2019, with an estimated 275 million women affected globally ^1,2^. Despite this burden, women have historically been underrepresented in cardiovascular research and clinical trials, resulting in limited understanding of sex-specific as well as gender-specific risk factors, atypical symptom presentations, and differential treatment outcomes ^1,2^. The persistent underrepresentation of women in cardiovascular research contributes to inequities in diagnosis, care, and health outcomes ^3^. Achieving equity in cardiovascular research therefore requires more than simply increasing women’s participation; it demands a systematic integration of sex and gender considerations throughout the entire research process—from study design and data collection to analysis and reporting.

The Canadian Institutes of Health Research (CIHR), Canada’s federal funding agency for health research and the largest funder of health research in the country, has progressively integrated sex- and gender-based analysis (SGBA) into its health research funding policies over the past two decades. Initial Policy and Guidelines introduced in 2006 encouraged applicants and reviewers to consider sex and gender in study design ^4^. In 2010, CIHR made SGBA mandatory in all grant applications by adding questions requiring applicants to specify whether and how sex (biological) and gender (socio-cultural) factors were addressed in their proposals ^4,5^. This milestone marked a fundamental policy shift toward embedding SGBA across the entire research cycle. To strengthen implementation, CIHR launched the SGBA in Research Action Plan in 2016-2017 and has since continued to promote systematic integration of sex and gender through ongoing initiatives ^4^. In 2018, CIHR further strengthened SGBA implementation at the evaluation stage by requiring peer reviewers to assess and rate the quality of sex and gender integration in each grant application, linking these assessments to funding decisions ^6,7^. The Institute of Gender and Health (IGH) Research Priority Plan 2024-2029 reaffirms CIHR’s long-term commitment to inclusive, intersectional health research and identifies women as a priority research population ^8^.

However, more than a decade after the SGBA mandate, the extent to which these policies have influenced actual funding allocation patterns remains unclear. Although CIHR’s multicomponent SGBA policy has led to measurable improvements in the reporting of sex and gender considerations by researchers at the research proposal stage ^6,9^, recent evidence indicates that these mandates have not substantially shifted the overall funding distribution toward women’s health research ^10^. Quantitative evaluations of how the SGBA mandate has influenced disease-specific investments remain scarce.

To address this gap, the present study used publicly available funding data from CIHR covering 2000 to 2024 to examine investment trends in cardiovascular research, with a focus on projects that incorporate sex or gender considerations or explicitly address women’s cardiovascular health. We classify all funded projects by the presence or absence of sex/gender terms and quantify changes in their investment share over time. To evaluate the potential impact of CIHR’s 2010 SGBA policy, we apply an interrupted time series (ITS) regression model to assess both immediate level changes and post-policy trend differences in women-related cardiovascular research funding. This study provides, to our knowledge, the first national-level quantitative evaluation of the SGBA policy’s effect on research funding patterns. Findings from this analysis offer critical evidence for funders and policymakers seeking to strengthen accountability and to advance sex and gender equity in Canadian health research.

## Methods Database

We analyzed the CIHR Grants and Awards datasets (2000-2024) to examine funding trends in cardiovascular research, with a particular focus on funding allocated to women-related research ^11^. These records correspond to Fiscal Years (FY) 2000-2001 through 2024-2025. Funding was measured using the CIHR variable *AmountPaidFY*, which reports the actual payments disbursed to each funded project per fiscal year. Multi-year grants therefore appear across multiple fiscal years according to their annual payments, without averaging or redistribution applied. This approach was applied consistently to both total cardiovascular research investment and women-related investment. This analysis included all direct research grants and operating funds but excluded awards primarily supporting trainee salaries and team/chair/training grants investments ^12^. Identification and classification of women-related cardiovascular research followed an inclusive definition detailed below.

### Search strategy

Search strategies for identifying cardiovascular research, sex and gender information are summarized in **Table 1**. All terms were searched in both English and French. For cardiovascular research identification, three fields were searched: title, keywords and research categories. To improve accuracy, “research categories” was used as the primary field because abstract-based searches may capture unrelated projects that mention “heart” or “stroke” incidentally.

**Table 1.** Summary of searching strategies.

| Identification goal | Searching fields | Searching terms |
| --- | --- | --- |
| Cardiovascular research | Title, keywords, research categories | Cardiovascular, cardiac, heart disease, heart attack, CVD, hypertension, coronary artery disease, peripheral artery disease, arrhythmia, microvascular angina, coronary syndrome, coronary microvascular dysfunction, coronary microvascular disease, INOCA, MINOCA, atrial fibrillation, angina pectoris, coronary, atherosclerosis, heart failure, myocardial infarction, myocardial ischemia, heart attack, coronary disease, STEMI, NSTEMI, cardiac infarction, ischemic heart disease, MINOCA, stroke, cerebrovascular disease, cerebrovascular accident, cerebrovascular apoplexy, brain infarction, cerebral ischemia, intracerebral hemorrhage |
| Sex and/or gender | Title, keywords, abstract | Sex, gender, non-binary, nonbinary, queer, two-spirit, 2S, hormone therapy, contraception |
| Female and/or women | Title, keywords, abstract | Female, women, maternal, menopause, gynecology, ovary, ovarian, uterus, uterine, cervix, fibroid, endometriosis, menstruation, obstetric, gestation, pregnant, pregnancy, lactating, postpartum, preeclampsia, amenorrhea, estrogen, progesterone |
| Male and/or men | Title, keywords, abstract | Male, men, testis, testicular, scrotum, penis, prostate, androgen, testosterone, epididymis, seminal vesicle, ejaculatory duct, sperm, spermatozoa, semen, prostate, testicular, penile, prostatectomy, varicocele, cryptorchidism, erectile dysfunction, andropause, hypogonadism, prostatitis, gynecomastia |

Sex and gender-related information were searched across title, keywords, and abstract fields using three categories: sex and/or gender terms (sex, gender, etc.), “female and/or women” terms (e.g., women, female, maternal, menopause, pregnancy, etc.), and “male and/or men terms (e.g., men, male, prostate, testosterone, etc.). The combinations of these indicators generated five categories of projects:

1. No sex or gender information: no relevant terms detected in any field;
2. Not specified: only “sex and/or gender” terms detected;
3. All: all three categories detected;
4. Only female and/or women: “sex and/or gender” and “female and/or women” terms detected, “male and/or men” absent;
5. Only male and/or men: “sex and/or gender” and “male and/or men” terms detected, “female and/or women” absent.

### Definition of women-related research

In this analysis, “women-related cardiovascular research” refers to projects that explicitly address females, women, sex, or gender dimensions of cardiovascular health. Projects in categories of “no sex or gender information” or “only male and/or men” were excluded because these projects were assumed not to meaningfully address sex or gender differences or were male or men-specific and therefore unlikely to contribute to women’s health improvement. More specifically, based on the search strategy summarized in **Table 1**, we included any project that met one or more of the following criteria:

1. Containing female or women-related terms (e.g., maternal, menopause, ovarian, uterine, pregnancy, postpartum, etc.);
2. Including sex or gender terms (e.g., sex, gender, hormone therapy, contraception, etc.) but excluding male or men-specific research; or
3. Targeting populations that self-identify as women, including cisgender and transgender women.

We adopted this inclusive definition recognizing that women’s cardiovascular health can be influenced by both biological sex and gendered social factors ^13^, and that research addressing sex or gender considerations that are not male or men specific may generate benefits for women, regardless of whether “women” were explicitly named as the study population. Therefore, both female/women-focused projects and sex/gender-focused projects were classified collectively as “women-related” cardiovascular research.

### Outcomes

The primary outcome was the annual proportion of CIHR cardiovascular research investment allocated to women-related research, defined as women-related fiscal-year payments divided by total CIHR cardiovascular research payments.

### Statistical analysis

We conducted an ITS analysis using segmented regression to evaluate the impact of CIHR’s two major SGBA policy interventions on the proportion of women-related cardiovascular research investment. The first interruption corresponds to the application-level SGBA policy introduced in 2010, when CIHR began requiring applicants to indicate whether and how sex and gender were considered in their research proposals. The second interruption represents the evaluation-level SGBA policy implemented in 2018, when peer reviewers were required to assess and rate the quality of sex and gender integration in each application. A quadratic term was included to capture potential non-linear acceleration in the post-2018 period.

The segmented regression model was specified as:

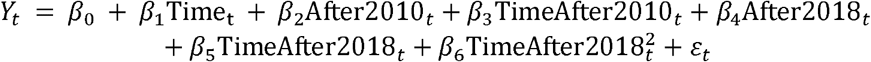

Where *Y*_*t*_is the proportion of total cardiovascular research funding allocated to women-related research in FY *t*. Time _t_is a continuous variable starting at 0 in FY 2000-2001 and increasing by 1 each subsequent FY. After2010_*t*_ is a dummy variable set to 0 before FY 2010-2011 and 1 thereafter. TimeAfter2010_*t*_ counts fiscal years since FY2010-2011 (0 before FY 2010-2011, 1 in FY 2010-2011, 2 in FY 2011-2012, etc.). After2018_*t*_ and TimeAfter2018_*t*_ are similarly defined for the second intervention in FY 2018-2019. 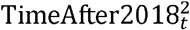 is the quadratic term, included to test for non-linear acceleration post-2018.

The coefficients are interpreted as follows: *β*_0_ estimates the baseline level of funding proportion at the start of the study period (FY 2000-2001).*β*_1_ is the linear trend prior to the 2010 application-level SGBA policy.*β*_2_ and *β*_4_ estimate the immediate level change in the funding level following the 2010 application-level and 2018 evaluation-level SGBA policy, respectively. *β*_3_ and *β*_5_ estimate the change in the linear slope following the 2010 application-level and 2018 evaluation-level SGBA policy, respectively.*β*_6_ captures for non-linear acceleration in the post-2018 period.

To address the observed autocorrelation in the model residuals, we used ordinary least squares (OLS) combined with Newey-West robust standard errors, which provide valid inference despite unspecified forms of autocorrelation and heteroskedasticity. A lag length of six was selected for the Newey-West correction based on inspection of the autocorrelation function plot of the initial OLS model’s residuals. All analyses were performed using R (version 4.5.2).

## Results

### Data screening and sex/gender representation

After screening the CIHR funding database, A total of 17,168 cardiovascular research records were identified for analysis (**Figure 1**). Each record represents a project-FY unit, as each CIHR project may have multiple records spanning its duration. Among the cardiovascular research grants, the majority (87.87%) did not include any sex or gender information, while 0.8% referred only to men (**Figure 2**). These two groups were excluded from subsequent analyses. Overall, between 2000 and 2024, 11.33% of annual CIHR cardiovascular research grants records met the criteria for women-related research, indicating that sex and/or gender considerations appeared in only a small proportion of cardiovascular research during this period.

**Figure 1.**
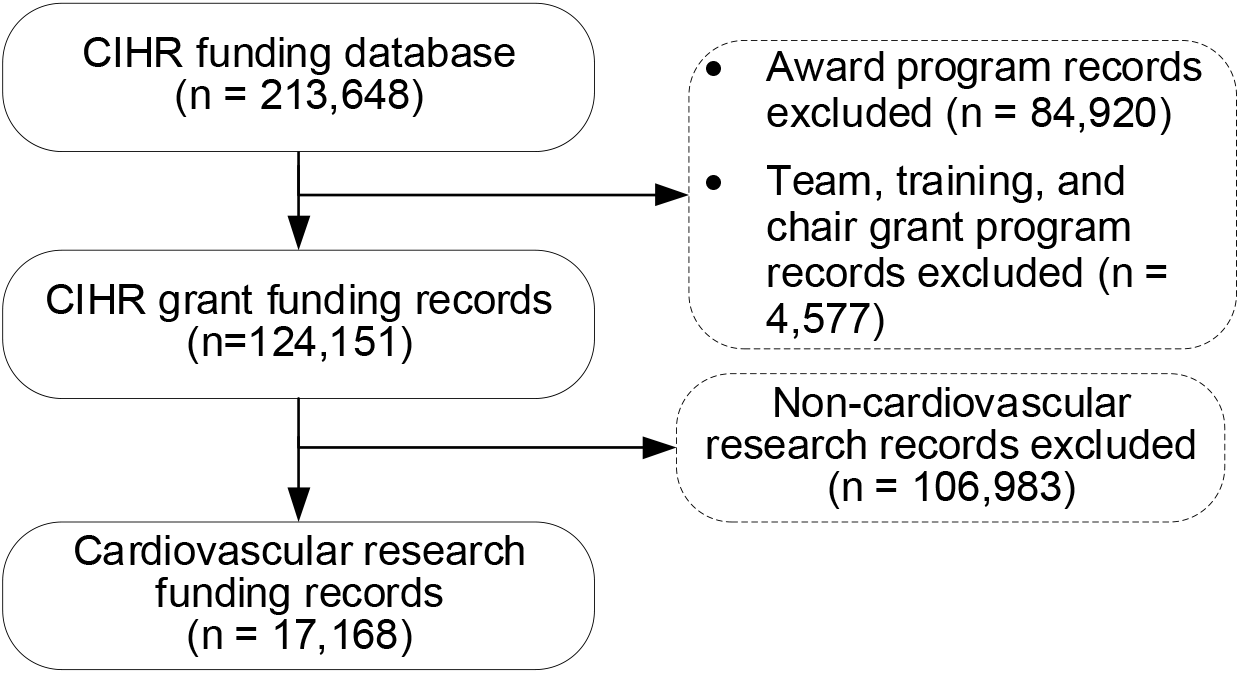
Screening flow of CIHR funding investment in cardiovascular research Notes: CIHR, Canadian Institutes of Health Research; n, represents the number of records, one programme may have several records across years.

**Figure 2.**
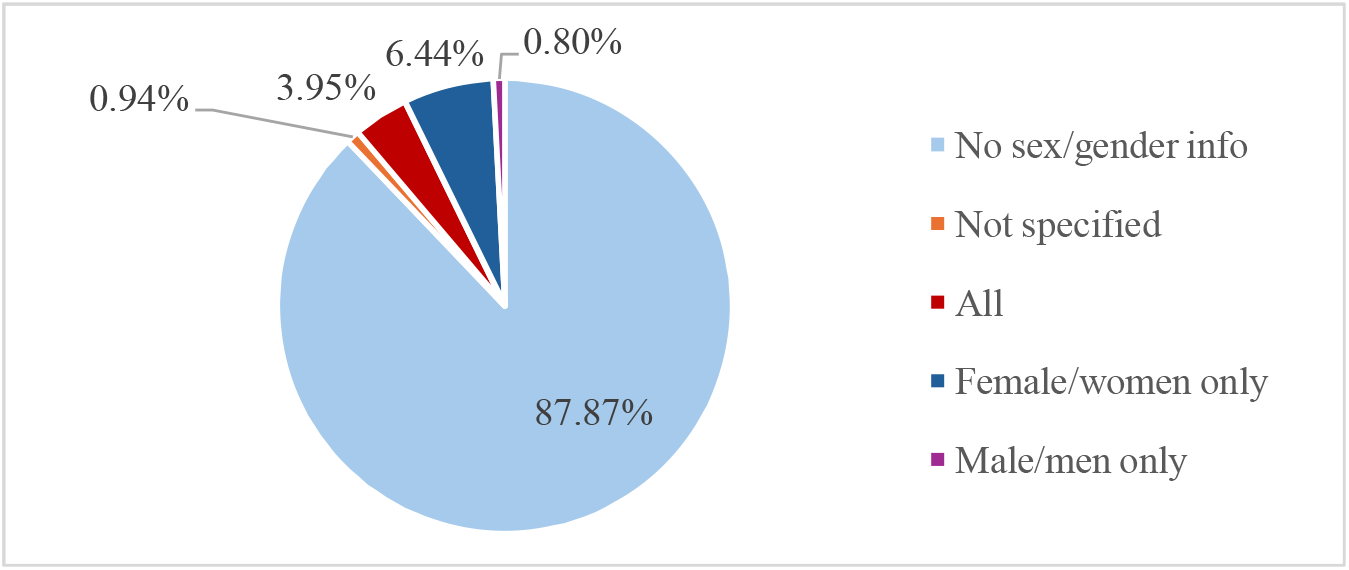
Sex and gender information groups and their proportions Note: The proportions were calculated by the number of records not the value of investments.

### Trends in CIHR cardiovascular research investment

Figure 3. illustrates annual trends in CIHR cardiovascular research funding by sex/gender relevance from 2000 to 2024. Overall, total cardiovascular research investment (blue bars) increased steadily from $40.59 million CAD in FY 2000-2001 to $122.88 million CAD in FY 2024-2025. Funding for women-related projects (red bars) rose from $1.30 million to $23.99 million CAD over the same period.

Despite this growth in absolute funding, the proportion of investment to women-related cardiovascular research remained modest. Women-related projects accounted for 3.21% of total cardiovascular research funding in FY 2000-2001, peaked at 19.52% in FY 2024-2025, and represented 11.85% of the total cardiovascular research cumulative funding disbursed across the 25-year period.

**Figure 3.**
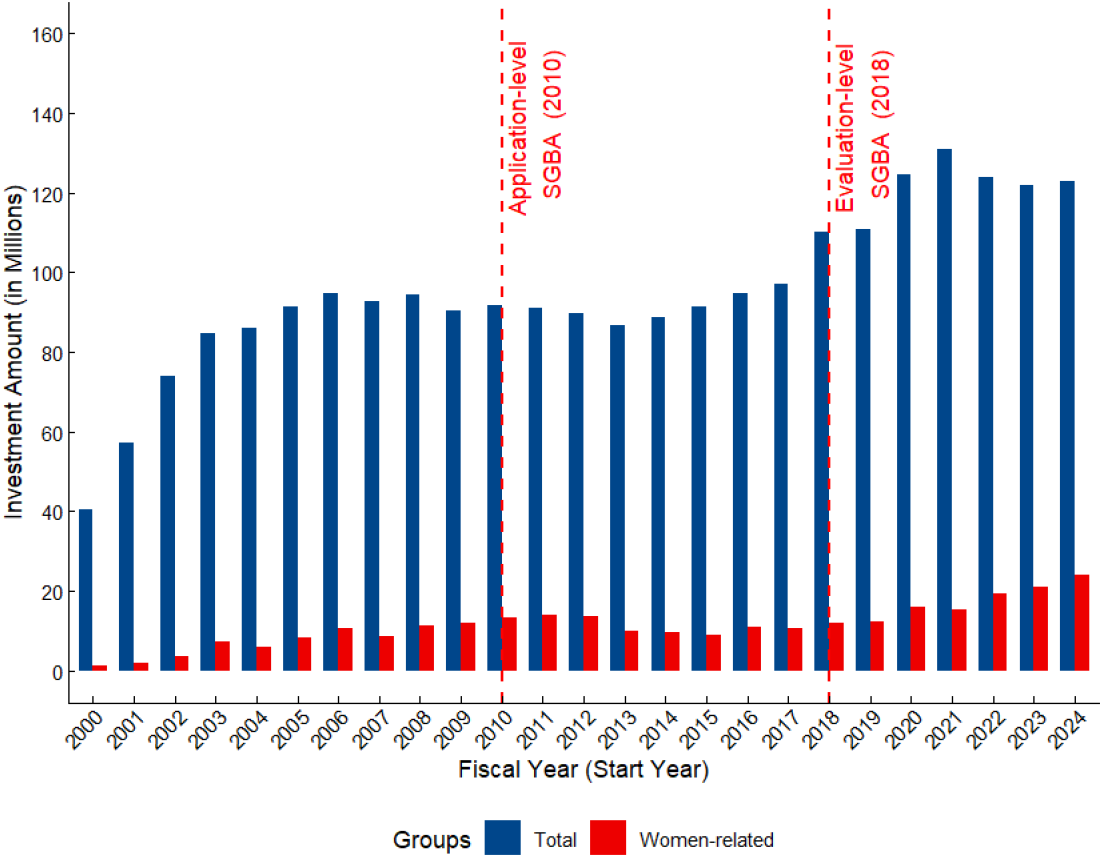
Annual trends in CIHR’s cardiovascular research investment and the share allocated to women-related research (2000-2024) Notes: CIHR, Canadian Institutes of Health Research; All amounts are expressed in current-year Canadian dollars.

### Segmented regression results

**Table 2** presents the segmented regression analysis of the proportion of CIHR cardiovascular research funding allocated to women-related research from 2000 to 2024. The estimated baseline proportion in 2000 was 2.2%, with a significant upward trend prior to 2010 (***β***_**1**_ = 0.0109, *P* < 0.001), indicating an average annual increase of about 1.1 percentage points before the policy implementation. Following the 2010 application-level SGBA policy, there was a modest immediate increase (***β***_**2**_ = 0.0260, *P* = 0.023), but the slope decreased significantly afterward (***β***_**3**_ = ™0.0181, *P* < 0.001), suggesting a temporary decline in growth. The 2018 evaluation-level SGBA policy did not lead to a statistically significant immediate change (***β***_**4**_ = 0.0099, *P* = 0.383) or a linear trend shift (***β***_**5**_ = 0.0029, *P* = 0.605). However, the quadratic term was positive and significant (***β***_**6**_ = 0.0024, *P* = 0.001), indicating a clear non-linear acceleration in women-related cardiovascular research investment in the most recent years.

**Table 2.**
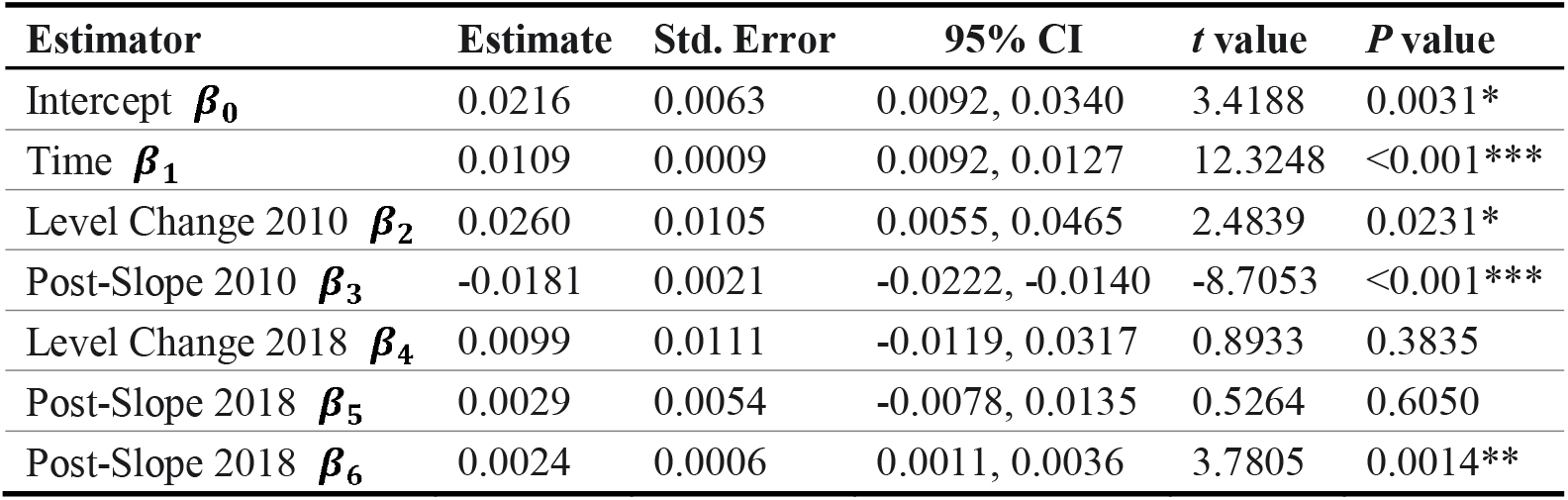
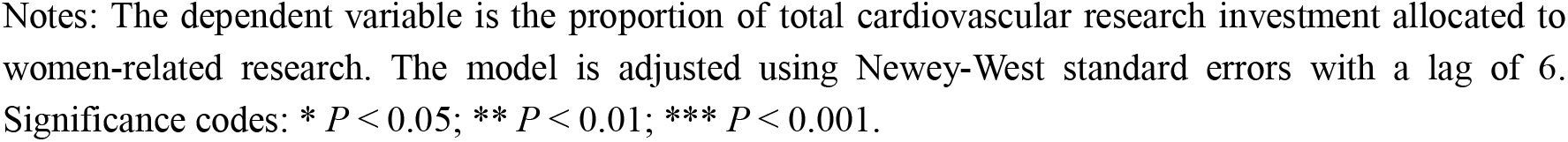
Segmented regression results for women’s cardiovascular research investment proportion (2000-2024)

| Estimator | Estimate | Std. Error | 95% CI | t value | P value |
| --- | --- | --- | --- | --- | --- |
| Intercept $\beta_0$ | 0.0216 | 0.0063 | 0.0092, 0.0340 | 3.4188 | 0.0031* |
| Time $\beta_1$ | 0.0109 | 0.0009 | 0.0092, 0.0127 | 12.3248 | <0.001*** |
| Level Change 2010 $\beta_2$ | 0.0260 | 0.0105 | 0.0055, 0.0465 | 2.4839 | 0.0231* |
| Post-Slope 2010 $\beta_3$ | -0.0181 | 0.0021 | -0.0222, -0.0140 | -8.7053 | <0.001*** |
| Level Change 2018 $\beta_4$ | 0.0099 | 0.0111 | -0.0119, 0.0317 | 0.8933 | 0.3835 |
| Post-Slope 2018 $\beta_5$ | 0.0029 | 0.0054 | -0.0078, 0.0135 | 0.5264 | 0.6050 |
| Post-Slope 2018 $\beta_6$ | 0.0024 | 0.0006 | 0.0011, 0.0036 | 3.7805 | 0.0014** |

Notes: The dependent variable is the proportion of total cardiovascular research investment allocated to women-related research. The model is adjusted using Newey-West standard errors with a lag of 6. Significance codes: \* $P < 0.05$ ; \*\* $P < 0.01$ ; \*\*\* $P < 0.001$ .

Figure 4. displays the observed and fitted trends in the proportion of women-related CIHR cardiovascular research funding. The observed data (blue line) show an increase before 2010, a period of decrease between 2011 and 2017, and a renewed rise after 2018. The fitted trend (red line) aligns well with these patterns, indicating a temporary decline following the 2010 policy introduction and a subsequent non-linear acceleration after the 2018 evaluation-level reinforcement.

## Discussion

### Main findings

Our analysis of CIHR cardiovascular research funding from 2000 to 2024 revealed that women-related cardiovascular research projects collectively accounted for 11.85% of the total funding disbursed over the entire period, representing a persistently modest share of overall cardiovascular research investment. Total cardiovascular research funding increased from $40.59 million CAD in FY 2000-2001 to $122.88 million CAD in FY 2024-2025, while investment in women-related cardiovascular research projects rose from $1.30 million CAD to $23.99 million CAD. Across the 25-year period, women-related projects accounted for an average of only 11.85% of total CIHR cardiovascular research investment. The proportion of women-related cardiovascular research investment followed three distinct phases: a steady rise before 2010, a marked decline between 2011 and 2017, and a renewed increase after 2018. Segmented regression analysis confirmed this temporal pattern, including a significant post-2018 acceleration captured by the quadratic term. These findings indicate that although investment in women-related research has increased in absolute terms, its overall share has progressed unevenly, with the most sustained growth only appearing in recent years.

**Figure 4.**
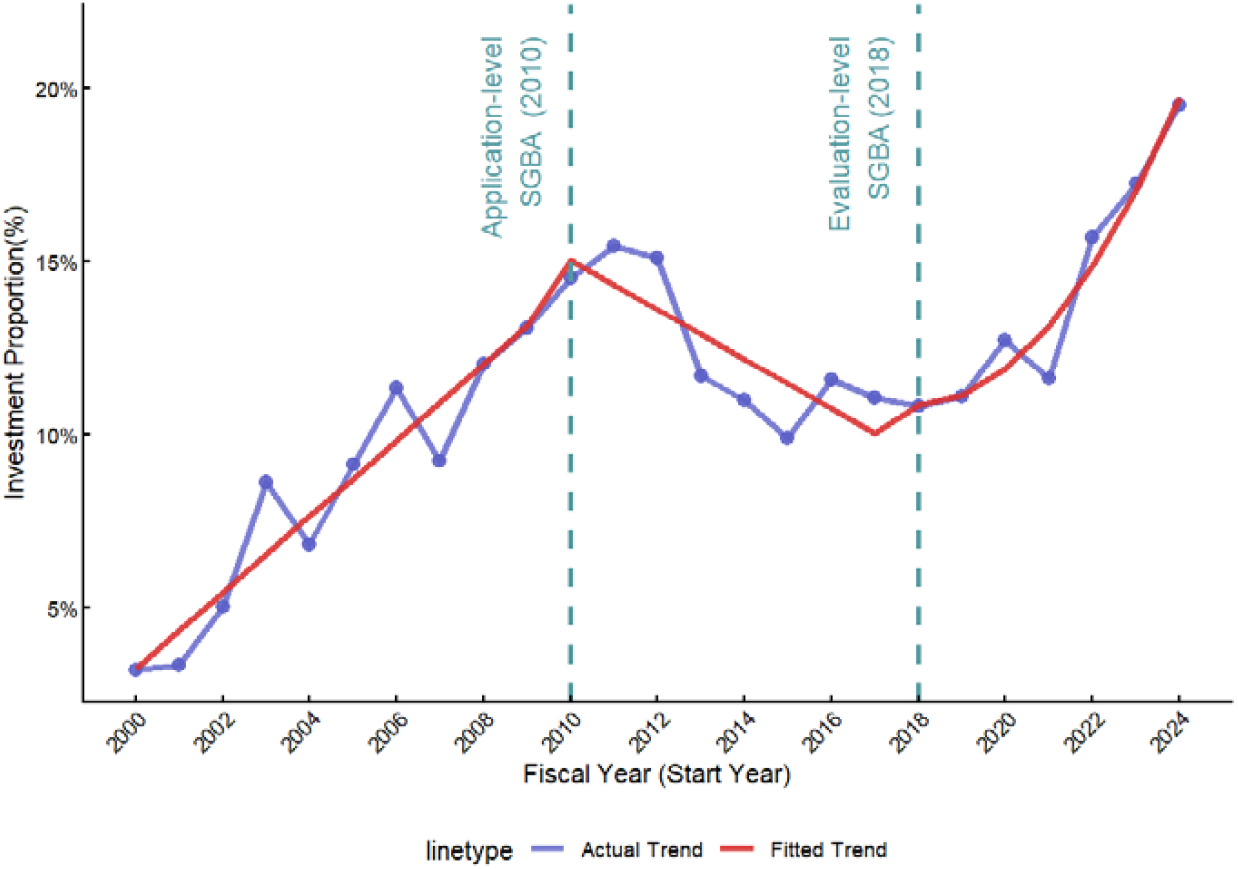
Observed and fitted trends in the proportion of women-related CIHR’s cardiovascular research investment (2000-2024) Note: CIHR, Canadian Institutes of Health Research.

### Literature contributions

This study refines existing terminology-based approaches by applying them within a single disease area where sex and gender differences are already well established. Like Stranges et al. (2023) and Gravelsins et al. (2025) ^5,10^, we identified women-related projects through funded projects’ titles, keywords, and abstracts, but we restricted the analysis to cardiovascular research, a field in which women’s differential symptoms, diagnostic experiences, and treatment responses are extensively documented ^1,2^. By calculating, for each FY, the proportion of total cardiovascular research funding directed to women-related projects, this study shifts the focus from the broad question of whether sex- or gender-related terms appear in CIHR-funded research to a more specific question: whether women receive a proportionate share of research investment in a disease area characterized by high disease burden and documented harms from inadequate sex- and gender-informed prevention, diagnosis, and care ^1,2,14^.

In addition, this study advances methodological approaches to analysing women-related research investment by incorporating fiscal-year funding data and applying time-series techniques capable of detecting structural changes. Previous CIHR-based analyses, such as Stranges et al. (2023) ^5^ and Gravelsins et al. (2025) ^10^, allocated the full value of multi-year grants to the competition year, which reflects planned commitments rather than the year-to-year flow of investment. Kim et al. (2024) examined the relationship between sex/gender-sensitive publications and grants using same-year counts, a design that does not account for the lag required for policies to influence funding decisions and research outputs ^15^. By analysing actual annual disbursements and using segmented regression with two interruption points and a quadratic term, the present study captures turning points and non-linear changes in women-related cardiovascular research investment that cross-sectional comparisons and competition-year allocations cannot reveal. This approach offers a more temporally sensitive framework for evaluating how research investment evolves over time and how it responds to major institutional interventions.

### Policy contributions

The pattern of women-related cardiovascular research investment, which first increased, then declined, and ultimately rose again after 2018, provides useful insight for strengthening existing policies. The steady increase before 2010 likely reflects growing awareness of sex and gender disparities in heart disease and early voluntary efforts to address them ^16,17^. The decline following the 2010 application-level SGBA requirement may suggest that merely requiring applicants to report sex and/or gender considerations had limited influence, consistent with earlier analyses ^5^. A complementary explanation is that SGBA content was increasingly placed within mandatory SGBA sections rather than in titles, keywords or abstracts after 2010, as indicated by internal application analyses and reviewer comments ^6,9^. This shift would reduce the appearance of women-related terminology in publicly available records even if research teams may have meaningfully considered these issues. The renewed rise after 2018 coincided with the introduction of SGBA guidance for reviewers and occurred during a period of growing national attention to women’s cardiovascular research in Canada ^6,18^. Although the CIHR-Heart & Stroke Foundation Initiative, launched in 2018, did not directly contribute to the funding amounts analyzed in this study because these awards were excluded, it formed part of the broader environment in which these trends occurred^18^. The trend observed in this study suggests that continued refinements to SGBA implementation may further enhance the visibility and prioritization of women-related cardiovascular research.

Although the recent increase is encouraging, women-related cardiovascular research investment has remained modest over the past 25 years, collectively accounting for approximately 11.85% of the total investment. Critically, the remaining 88% of projects did not explicitly report sex- or gender-related considerations, making it difficult to determine whether the funded research would meaningfully address women’s cardiovascular health needs. Given the well-documented historical underrepresentation of women in cardiovascular research and the long-standing gaps in sex- and/or gender-specific evidence, the persistently modest share of women-related projects suggests that important knowledge needs remain insufficiently addressed ^3,14^. Moreover, terminology-based identification captures only the most basic layer of women-related research.

Projects mentioning women or sex or gender in their titles, keywords, and abstracts vary widely in substantive SGBA integration. Many simply include women in the study population without analyzing sex or gender differences. Recent analysis from McKinsey indicates that only about half of evaluated interventions report sex-disaggregated data, highlighting a persistent gap between inclusion and meaningful sex-specific evidence ^19^. Strengthening women-related cardiovascular research is not only important for health equity but also economically justified. Analyses from RAND and McKinsey show that increasing investment in women’s health research generates substantial returns, including productivity gains, reduced healthcare costs, and broader societal benefits ^20-23^. These findings underscore the value and importance of sustained growth in women-related cardiovascular research.

International experience shows substantial variation in the timing and strength of policies designed to integrate sex and gender into health research systems. In the United States, the NIH’s Sex as a Biological Variable (SABV) policy (2016) mandates consideration of sex in study design, analysis, and interpretation and embeds these requirements directly into peer review, representing one of the strongest and earliest institutional approaches ^24,25^. By contrast, the United Kingdom’s recent initiatives reflect a more moderate level of enforcement. The *Medical Research Council (MRC) Guidance for the Inclusion of Sex in Experimental Design* (2022) requires investigators who propose single-sex studies to provide strong scientific justification, and such justification is reviewed as part of the peer-review process, although it is not embedded as a scored review criterion ^26^. The Women’s Health Strategy for England (2022) further highlights the importance of considering sex differences in publicly funded research, but functions primarily as a strategic policy directive rather than a mandatory review requirement ^27^. Australia has also advanced its policy framework in recent years. The 2023 national *Statement on Sex, Gender, Variations of Sex Characteristics and Sexual Orientation in Health and Medical Research* encourages the systematic consideration of sex and gender across study design, recruitment, analysis, and reporting ^28^. However, these policy expectations in Australia remain guidance-based and have not yet been formalized as mandatory or scored review criteria.

Compared with these jurisdictions, Canada adopted sex- and gender-based analysis earlier, introducing SGBA requirements at the application level in 2010 and subsequently strengthening them by incorporating SGBA into peer-review guidance in 2018. The post-2018 increase observed in women-related cardiovascular research investment in this study is consistent with evidence from the United States ^15,24,25^, suggesting that reviewer-level assessment mechanisms are more likely than application-level reporting to influence research priorities and funding allocation.

Building on the SGBA foundation, further practical steps could enhance the integration of women-related evidence. A simplified tiered SGBA indicator that distinguishes inclusion in the study population, disaggregated analyses, sex and gender-informed interpretation, and mechanism-focused investigation would allow more meaningful monitoring while maintaining a manageable reporting burden for researchers. Evidence from CIHR’s SGBA reforms shows that reviewer training and the inclusion of SGBA quality criteria in scoring systems have strengthened the link between SGBA considerations and funding decisions, addressing the gap between improved SGBA documentation and actual research priorities ^6^. Targeted funding programs also offer a constructive way to advance women-related cardiovascular science.

Initiatives, such as the CIHR-Heart & Stroke Foundation women’s heart and brain health program and the newly launched national Women’s Heart and Brain Health Networks, show that directed investment can build research capacity and encourage coordinated research activities ^18,29^. Finally, establishing a longitudinal, open monitoring system that links annual funding patterns with downstream outcomes—including sex-disaggregated publications, clinical guideline updates, and knowledge translation—would help evaluate the long-term impact of investment. Such a system could also incorporate an accountability indicator assessing whether funded projects ultimately carried out the SGBA analyses they committed to in their original grant applications.

### Limitations

First, women-related cardiovascular research was identified using terminology in funded titles, keywords, and abstracts. This may underestimate women-related research if relevant content was placed in SGBA-specific sections of the application or incorporated only at the publication stage, and it may overestimate substantive integration when terminology does not translate into sex-disaggregated analyses or mechanism-focused. Thus, terminology-based identification reflects only the visibility of women-related content and does not indicate whether a study meaningfully advances cardiovascular health outcomes for women. Second, although the time-series design captures temporal shifts, it cannot attribute changes to SGBA policies alone. Observed trends may also reflect shifts in CIHR strategic priorities, targeted federal initiatives, or broader developments in the cardiovascular research landscape.

## Conclusions

This study provides the first long-term assessment of CIHR’s investment in women-related cardiovascular research and shows recent increases in the share of funding allocated to women-related cardiovascular research. Although women-related cardiovascular research funding increased after 2018, it has remained modest over the past 25 years, and the vast majority of cardiovascular research projects (88%) did not explicitly report sex- or gender-related information. The temporal patterns observed—particularly the limited effect of the 2010 application-level SGBA requirement and the renewed rise following the 2018 reviewer-level guidance—highlight the importance of embedding sex and gender considerations within peer-review processes rather than relying on applicants’ statements at the proposal stage. Strengthening the implementation of SGBA, expanding targeted funding initiatives, and establishing ongoing monitoring of sex-disaggregated outputs would support a more equitable and evidence-informed allocation of research funding. Continued progress in these areas is essential to closing longstanding gaps in women’s cardiovascular research and achieving better health outcomes for women in Canada.

## Data Availability

The data analyzed in this study are publicly available from the Canadian Institutes of Health Research (CIHR) Grants and Awards datasets on the Government of Canada Open Data Portal.

https://open.canada.ca/data/en/dataset/49edb1d7-5cb4-4fa7-897c-515d1aad5da3

